# Single-Cell and TCR Profiling across Tissues Reveals GZMK^+^CD8^+^ T Cells as Drivers of Fibrosis in IPF

**DOI:** 10.64898/2025.12.20.25342718

**Authors:** Bingqing Yue, Shuliang Jing, Qiaxuan Li, Guoyu Xie, Juan Chen, Tao Yan, Ganggui Zhu, Bin Huang, Jin Zhao, Xiucheng Yang, Jian Huang, Pan Yin, Qifeng Yao, Huijing Yang, Fei Gao, Jun Yang, Man Huang, Jingyu Chen

**Affiliations:** Department of Lung Transplantation, The Second Affiliated Hospital Zhejiang University School of Medicine, 1511 Jianghong Road, Binjiang District, Hangzhou 310051, Zhejiang Province, China; Key Laboratory of Multiple Organ Failure (Zhejiang University), Ministry of Education, Hangzhou 310051, Zhejiang, China; Department of Physiology, Zhejiang University School of Medicine, State Key Laboratory of Transvascular Implantation Devices, 866 Yuhangtang Road, Hangzhou 310058, Zhejiang Province, China; Department of General Intensive Care Unit, The Second Affiliated Hospital of Zhejiang University School of Medicine, Hangzhou 310051, Zhejiang Province, China; Department of Emergency, Wuxi People’s Hospital affiliated to Nanjing Medical University, Jiangsu Province 214000, China; Lung Transplant Center, Wuxi People’s Hospital affiliated to Nanjing Medical University, Jiangsu Province 214000, China

**Keywords:** GZMK, TGF-β1 signalling, ALK4, Idiopathic pulmonary fibrosis

## Abstract

Idiopathic pulmonary fibrosis (IPF) is a fatal interstitial lung disease with limited therapies and poorly defined cross-tissue immune mechanisms. We performed single-cell RNA sequencing and TCR profiling of paired lung, lymph node, and peripheral blood samples from patients with IPF, combined with functional coculture assays and mouse model. We identified GZMK-high CD8⁺ T cells enriched in fibrotic lungs, displaying inflammatory but low-cytotoxic features. TCR and trajectory analyses indicated that these cells originate from lymph-node CD8_HSPA1A cells and migrate to the lung. Higher GZMK⁺CD8⁺ T-cell levels correlated with impaired lung function and worse outcomes. Mechanistically, GZMK-overexpressing CD8⁺ T cells promoted fibroblast-to-myofibroblast differentiation and proliferation through TGF-β1 signalling, while GZMK inhibition reduced fibroblast activation and collagen deposition. Pharmacological blockade of TGFβR1/ALK4 with TEW suppressed fibroblast activation in vitro and significantly attenuated pulmonary fibrosis in vivo. Together, these findings identify a lymph node–to–lung migration axis of GZMK⁺CD8⁺ T cells that drives fibroblast activation through TGFβR1/ALK4 signalling, highlighting GZMK as a potential biomarker and therapeutic target in IPF.

## Introduction

Idiopathic pulmonary fibrosis (IPF) is a progressive, fatal interstitial lung disease of unknown aetiology characterized by relentless fibrotic remodelling of the pulmonary parenchyma, ultimately culminating in end-stage respiratory failure^1^. The median survival following diagnosis is approximately 2–4 years, with a 5-year survival rate less than 20%, underscoring the urgent unmet clinical need for novel therapeutic approaches^2,3^. Histologically, the hallmark features of IPF include the aberrant proliferation and activation of fibroblasts, their transition into α-smooth muscle actin (α-SMA)-expressing myofibroblasts, and excessive extracellular matrix (ECM) deposition, culminating in irreversible fibrotic lesions termed “fibroblastic foci”^4^.

Recent studies have increasingly highlighted the critical role of immune regulation in IPF pathogenesis^5^. Mediastinal lymph node enlargement (MLNE), which is detectable by computed tomography (CT), has been reported in 40–80% of IPF patients^6,7^. MLNE is correlated with higher total lung fibrosis scores, reduced diffusing capacity for carbon monoxide (DLCO), and poorer prognoses compared to patients without lymphadenopathy, suggesting its potential utility as a predictive biomarker for disease progression^7–9^. The precise biological mechanisms underlying MLNE in IPF remain undefined, but this phenomenon likely reflects heightened systemic immune activation that is integral to pulmonary pathology. In recent years, single-cell RNA sequencing (scRNA-seq) has significantly advanced our understanding of the pathogenesis of IPF^10,11^. However, to date, no studies have performed single-cell transcriptomic profiling of enlarged lymph nodes in IPF patients, nor have they included matched analyses of peripheral blood and lung tissue samples. Therefore, elucidating the distinctive lymphocyte characteristics and immune microenvironments across diverse tissue compartments in IPF with combined sequencing techniques is imperative for the development of effective therapeutic strategies.

Granzyme K (GZMK) is a serine protease secreted by lymphocytes that induces inflammatory cell death or promotes the release of proinflammatory cytokines through noncanonical pathways^12^. In contrast to the classical cytotoxic molecule GZMB, GZMK is highly expressed in chronic inflammatory microenvironments and is proposed to function as an immunological mediator that both sustains cytotoxic immune responses and amplifies inflammatory signalling cascades^13^. Recent studies in the contexts of cancer and chronic diseases have demonstrated that GZMK⁺CD8⁺ T cells exhibit distinct chemotactic and cytokine secretion profiles^14,15^. Their activation can be directly induced by cytokines, providing a mechanistic rationale for their involvement in the pathogenesis of fibrosis.

Current first-line pharmacological treatments for IPF include pirfenidone and nintedanib^16^. Pirfenidone mitigates fibrosis progression primarily by inhibiting the expression of TGF-β1 and proinflammatory cytokines, whereas nintedanib is a multitarget tyrosine kinase inhibitor that blocks key signalling pathways such as PDGFR, FGFR, and VEGFR, thereby suppressing fibroblast proliferation and angiogenesis and slowing lung function decline^17,18^. However, neither agent can reverse established fibrosis or significantly improve overall survival, and both are associated with a certain degree of adverse effects, including gastrointestinal symptoms and photosensitivity, with limited target specificity^19,20^. Emerging therapeutic strategies targeting the TGF-β pathway, including agents such as αvβ6 and αvβ1 integrin inhibitors, are gaining attention^21^. However, these agents remain largely in the early stage of clinical research^22^. To date, no TGF-β receptor-targeted inhibitor has been specifically applied to IPF therapy. TEW-7197, also known as EW-7197 or vactosertib, is a highly potent, orally bioavailable small-molecule inhibitor of TGF-β type I receptor kinases ALK4 (ACVR1B) and ALK5 (TGFBR1). By occupying the ATP-binding pocket of these receptors, TEW-7197 blocks ligand-induced phosphorylation of SMAD2/3 and thereby suppresses canonical TGF-β/activin signaling. This blockade reduces transcription of TGF-β target genes involved in fibrosis, epithelial–mesenchymal transition, and tumor invasion, while also modulating the tumor microenvironment by alleviating TGF-β–mediated immunosuppression and potentially enhancing anti-tumor immunity^23–25^.

In this study, we performed single-cell RNA sequencing on paired lung parenchyma, peripheral blood, and mediastinal lymph node samples from IPF patients. We identified a GZMK⁺CD8⁺ T-cell subset that was selectively enriched in fibrotic lung tissue and characterized by a distinct inflammatory profile with relatively low cytotoxic potential. T cell receptor (TCR) sequencing further revealed a lymph-node–enriched CD8⁺ T-cell population (CD8_HSPA1A), defined by high expression of the stress-response chaperone HSPA1A and transcriptional features of recently activated effector memory cells, which represents a precursor state that gives rise to the pathogenic CD8_GZMK cells detected in lung tissues. Through in vivo and in vitro experiments, we demonstrated that GZMK⁺CD8⁺ T cells directly drive fibroblast-to-myofibroblast transition via the TGF-β1 signalling axis. Among IPF patients, those with higher GZMK⁺CD8⁺ T-cell scores had worse prognoses, suggesting GZMK as a potential diagnostic biomarker. TEW inhibited TGFβR1/ALK4 activity and effectively ameliorated bleomycin (BLM)-induced pulmonary fibrosis, highlighting a promising therapeutic approach.

## Results

### Landscape view of cell composition in lung tissues, lymph nodes, and peripheral blood in patients with IPF at single-cell resolution

To comprehensively characterize the immune microenvironment in idiopathic pulmonary fibrosis (IPF), we performed single-cell RNA sequencing (scRNA-seq) and TCR repertoire profiling on 11 samples from four IPF patients. These samples included matched lung parenchyma (PT), mediastinal lymph nodes (LN), and peripheral blood (PB) (**Figure 1A**). After stringent quality control filtering, doublet exclusion, and normalization, a total of 83,319 high-quality single cells were retained for downstream analysis.

**Figure 1.**
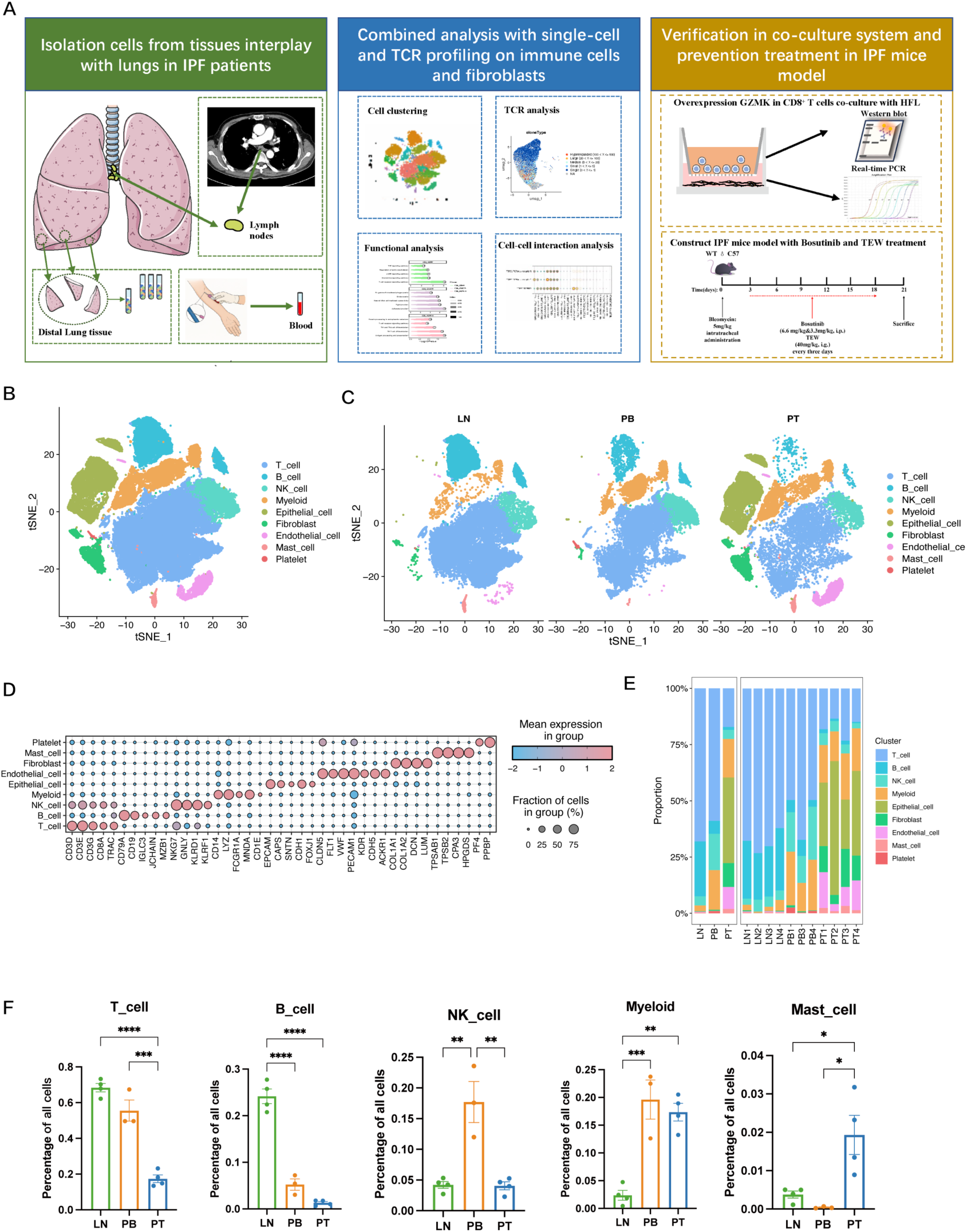
Single-cell landscape of immune and stromal cell composition in IPF lungs, lymph nodes, and peripheral blood. **A.** Overall study design diagram with 11 IPF samples constructed by using the 10X Genomics Platform. **B.** t-SNE plot showing 9 clusters of 83,319 cells. Each dot represents an individual cell, coloured according to its assigned cell type on the basis of transcriptional signatures. **C.** Dot plot showing marker gene expression for each cell type, with colour intensity indicating average expression levels and dot size representing the proportion of cells expressing each marker gene, as shown in the scale bar on the right. **D.** t-SNE plots showing the distribution of major cell populations across different sample sites, including LNs, PB, and PTs, from IPF patients. **E.** The left panel shows the proportional distribution of cell types across different tissue categories, including LN, PB, and PT samples. The right panel illustrates the cell type composition within individual samples from these tissue categories. **F.** Bar plot showing the percentages of T cells, B cells, NK cells, myeloid cells, and mast cells across the LN, PB, and PT compartments in IPF patients. The data are presented as the mean ± standard error of the mean. Statistical significance was determined using a one-way ANOVA with multiple comparisons. *P<0.05, **P<0.01, ***P<0.001, ****P<0.0001.

Unsupervised clustering and t-distributed stochastic neighbour embedding (t-SNE) visualization revealed nine major cell populations based on classical marker gene expression, including T cells, B cells, natural killer (NK) cells, myeloid cells, epithelial cells, endothelial cells, fibroblasts, mast cells, and platelets (**Figure 1B, Supplementary Figure 1A**). t-SNE plots stratified by sample origin including PT, LN, and PB revealed distinct tissue-specific clustering patterns (**Figure 1C**). The marker genes defining each cluster are illustrated in **Figure 1D** (see also **Supplementary Figure 1C**).

To elucidate the cellular composition across the PT, LN, and PB samples of IPF patients, we analysed the tissue-specific distributions of cell types (**Figure 1E and 1F, Supplementary Figure 1B**). The LN samples have enriched in T and B lymphocytes, which is consistent with their immune surveillance function. PB samples contained relatively high proportions of T and NK cells. PT samples showed prominent stromal cell components and immune cell infiltration characterized by a substantial proportion of T cells and myeloid cells, highlighting the immunologically active microenvironment within IPF lung tissues. Consequently, we focused on these immune cell subsets to explore their roles in IPF progression and differentiation across tissue compartments.

### Lung-Enriched CD8_GZMK T Cells Exhibit Inflammatory and Fibrogenic Signatures in IPF

T cells constitute pivotal components of the IPF microenvironment and are closely associated with disease progression^26,27^. Through unsupervised clustering analysis of 40,635 cells across all the samples, we identified five CD4^+^ T-cell clusters, four CD8^+^ T-cell clusters, one NKT cell cluster, and one MAIT cell cluster (**Figure 2A, Supplementary Figure 2A–C**). Tissue distribution analysis revealed compartment-specific enrichment of T-cell subsets, with notable differences in CD8⁺ T-cell composition across tissues (**Figure 2A**).

**Figure 2.**
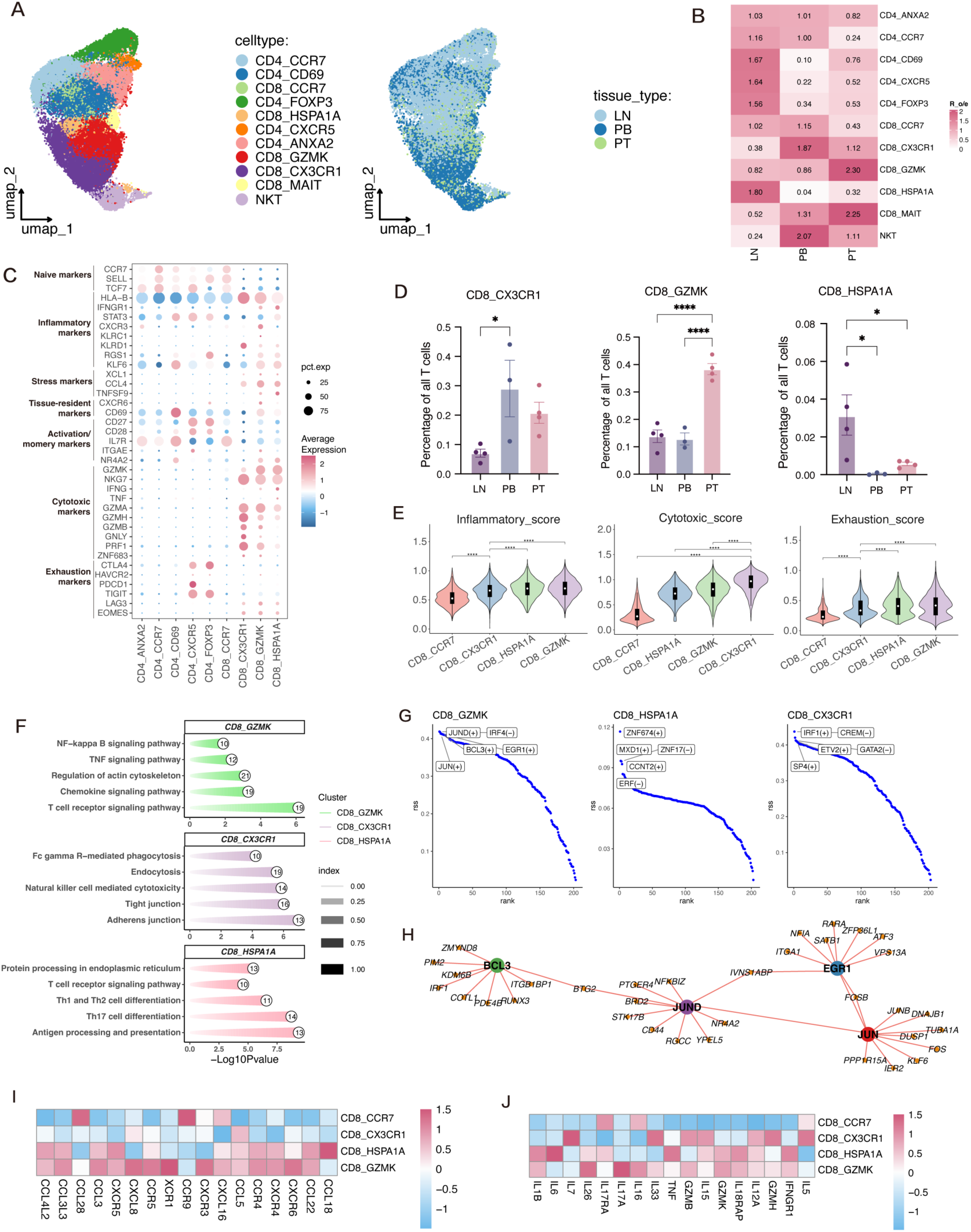
Proinflammatory GZMK^+^CD8^+^ T cells are enriched in IPF lung tissues. **A.** UMAP plot of T cells from IPF patients, with clustering into 11 distinct clusters, each represented by a different colour (left) and their distribution across different sample sites, including LN, PB, and PT (right). **B.** Heatmap displaying the tissue preference of each T-cell across different tissue compartments in IPF patients analysed by Ro/e. **C.** Dot plot showing the expression of functional marker genes across CD8^+^ T-cell subsets. **D.** Bar plot showing the percentages of CD8_ GZMK (GZMK^+^CD8^+^ T cells), CD8_ HSPA1A, and CD8_ CX3CR1 cells across the LN, PB, and PT compartments in IPF patients. Data are presented as mean ± standard error of the mean (SEM). Statistical significance was determined using a one-way ANOVA with multiple comparisons. *P<0.05, ***P<0.001, ****P<0.0001. **E.** Violin plot showing the inflammatory score, cytotoxic score, and exhaustion score across CD8⁺ T-cell subsets. **F.** KEGG pathway enrichment results for the CD8_ GZMK, CD8_ HSPA1A, and CD8_ CX3CR1 subsets based on the top genes from each cluster. **G.** SCENIC regulatory-signature ranking curves for CD8_GZMK, CD8_HSPA1A, and CD8_CX3CR1; leading-edge transcription factors are labelled. **H.** SCENIC-derived transcription factor network linking the four most influential regulons in the CD8_GZMK subset (BCL3, JUND, JUN, and EGR1) to their top target genes. **I.** Heatmap showing the expression of chemokine and chemokine-receptor genes across CD8^+^ T-cell subsets. **J.** Heatmap displaying the expression of proinflammatory cytokine molecules in CD8^+^ T-cell subsets.

Among these, CD8_CCR7 cells represent naïve T cells with high *CCR7*, *SELL*, and *TCF7* expression. CD8_CX3CR1 cells expressed elevated levels of *GZMH, KLRG1, CX3CR1,* and *PRF1*, corresponding to classical effector T cells, whereas the CD8_HSPA1A and CD8_GZMK subsets presented transcriptional signatures of effector memory (Tem) cells with distinct functional features (**Supplementary Figure 2B**). Notably, CD8_HSPA1A exhibited higher expression of heat shock protein family genes. Proportional analysis revealed that the CD8_GZMK subset was markedly enriched in PT samples, while CD8_CX3CR1 cells were more abundant in PB samples, and CD8_HSPA1A cells preferentially localized to LN samples (**Figure 2B and 2D, Supplementary Figure 2D**). Among CD8⁺ T-cell subsets, CD8_CX3CR1 cells showed the highest cytotoxicity scores, whereas CD8_GZMK cells had lower cytotoxicity scores than CD8_CX3CR1 cells but higher than naive CCR7⁺ CD8 T cells, together with the highest inflammatory and exhaustion scores. (**Figure 2C and 2E; Supplementary Figure 2E and 2F**).

To explore the functional roles of distinct CD8⁺ T-cell subsets in IPF, we performed pathway enrichment analysis on the differentially expressed genes in each cluster **(Figure 2F)**. The CD8_GZMK population, enriched in fibrotic lung tissues, showed significant upregulation of pro-inflammatory pathways including TNF signalling, chemokine signalling, and T cell receptor signalling. In contrast, CD8_CX3CR1 cells were enriched for cytotoxic immune response pathways, while CD8_HSPA1A cells demonstrated upregulation of antigen presentation and immune modulation pathways. Furthermore, Regulon analysis using SCENIC revealed that CD8_GZMK cells were transcriptionally driven by proinflammatory and immediate-early transcription factors, including *JUN*, *JUND*, *BCL3*, and *EGR1* **(Figure 2G)**. These TFs are known to regulate T-cell activation, cytokine production, and tissue remodelling. To further explore transcriptional regulation, we constructed a TF–target gene interaction network **(Figure 2H)**, identifying *JUN*, *JUND*, *BCL3*, and *EGR1* as central hub regulators connected to fibrosis- and inflammation-associated genes such as *PTGER4*, *NR4A2*, *CD44*, and *FOS*. This network suggests that CD8_GZMK cells may sustain chronic immune activation via a tightly coordinated regulatory program.

We further examined the expression of chemokines, chemokine receptors, and inflammatory cytokines across clusters (**Figure 2I and 2J**). CD8_GZMK cells exhibited upregulated expression of several chemokine receptors, including *CXCR4*, *CCR4*, and *CXCR6*, as well as chemokines such as *CCL3L3*, facilitating immune cell recruitment to inflamed tissues (**Figure 2I**). Additionally, CD8_GZMK cells exhibited marked upregulation of proinflammatory cytokines such as *IL17A*, *IL26*, and *IL1B* (**Figure 2J**). Notably, unlike the CD8_CX3CR1 subset, which showed high expression of classical cytotoxic genes such as *GZMB* and *PRF1*, CD8_GZMK cells exhibited a hybrid phenotype characterized by inflammatory cytokine production with limited cytolytic potential. Together, these results characterize CD8_GZMK cells as a low-cytotoxicity, inflammation-skewed effector memory CD8⁺ T-cell population enriched in the fibrotic lung, which may contribute to persistent immune activation and immune-mediated fibrogenesis in IPF.

### TCR analysis revealed a close relationship between CD8_GZMK and CD8_HSPA1A cells

To investigate the clonal architecture and developmental trajectories of T-cell subsets in IPF, we performed TCR sequencing and successfully recovered paired TCR α and β chains in 32,125 T cells across all the samples. Notably, compared with CD4⁺ T cells, CD8⁺ T cells exhibited a substantially greater degree of clonal expansion, suggesting that antigen-driven proliferation occurred within the fibrotic lung microenvironment (**Figure 3A and 3B)**. Among tissues, T cells from PT samples demonstrated the highest proportion of expanded clones, in contrast to the more quiescent profiles observed in PB and LN samples, indicating robust local immune activation at the site of fibrosis. Further analysis of T-cell clonal expansion in IPF revealed prominent clonal amplification within the CD8_CX3CR1, CD8_GZMK, and CD8_HSPA1A subsets, highlighting sustained or highly active immune responses within the local microenvironment (**Figure 3C**).

**Figure 3.**
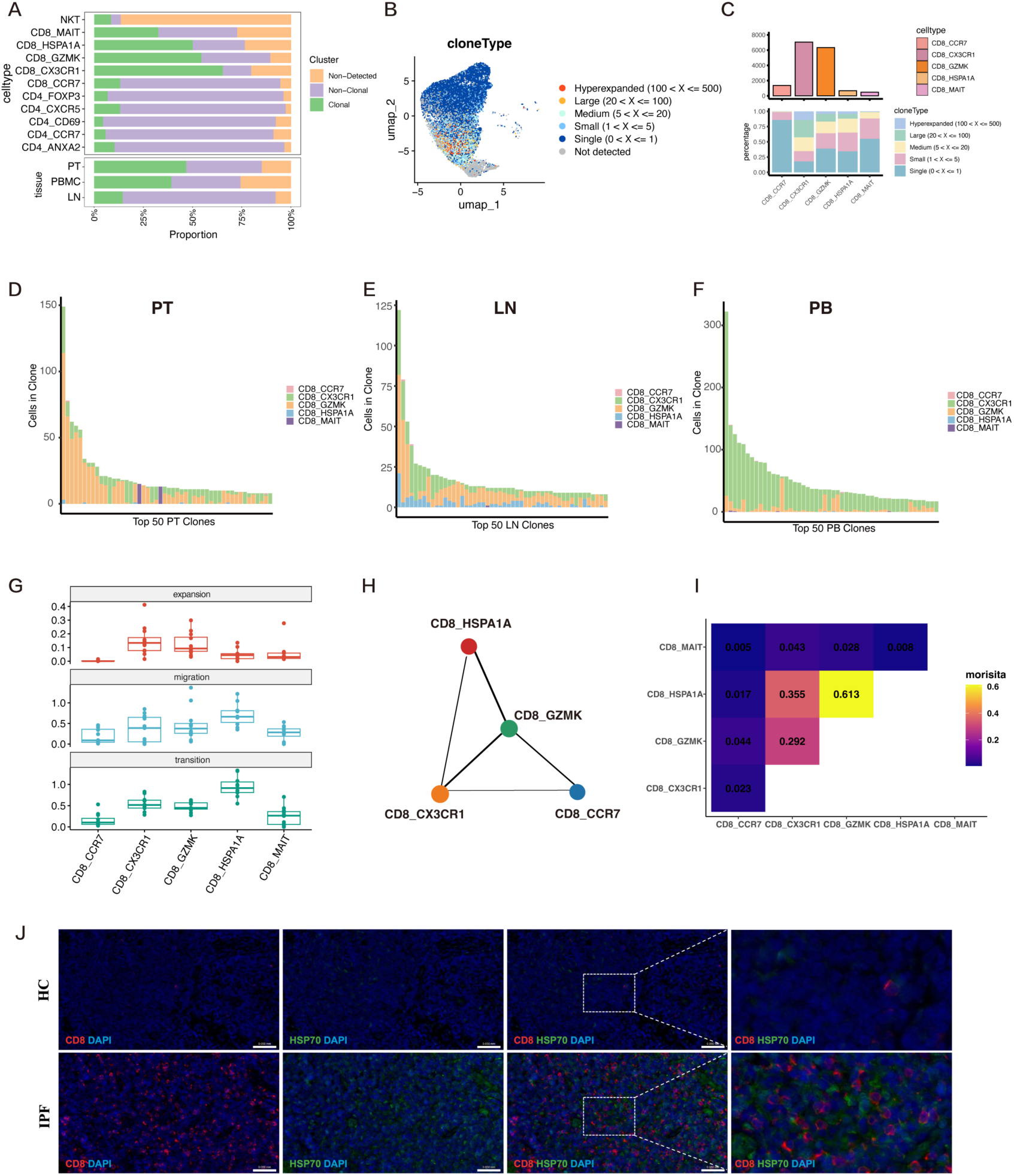
CD8_HSPA1A T cells in lymph nodes show clonal expansion with lung CD8_GZMK T cells in IPF. **A.** Proportions of clonal, non-clonal, and non-detected T cells across different subsets and tissues in IPF. **B.** UMAP visualization displaying the distribution of T-cell clonotypes on the basis of their clone sizes. **C.** Numbers and clonal expansion patterns of major CD8⁺ T-cell subsets in IPF. **D‒F.** Bar plots showing the sizes and cluster composition of the top 50 clones, along with the clone size distributions for the lymph nodes (D), peripheral blood (E), and pulmonary tissues (F). **G.** Clonal expansion, migration, and transition potential of CD8^+^ T cells assessed using STARTRAC indices. **H.** PAGA analysis illustrating CD8^+^ T-cell clusters, where each dot corresponds to a specific T-cell cluster. **I.** Heatmap showing the clonal overlap between CD8^+^ T-cell subsets derived from the TCR analysis. **J.** Representative images of multiplex immunofluorescence staining for HSPA1A^+^CD8^+^ T cells in the lymph nodes of patients with IPF and healthy donors. scale bar, 50 μm. n = 3 individual patient samples were examined independently.

To further delineate tissue-specific clonal dynamics, we profiled the top 50 expanded clones in each compartment. In PT, the top expanded clones were overwhelmingly dominated by the CD8_GZMK subset, with minimal contributions from other T-cell subsets (**Figure 3D**). This striking enrichment suggests that CD8_GZMK cells are the predominant antigen-experienced population at the site of fibrosis and may directly contribute to local inflammation and tissue remodelling in IPF. In the LN, the clones were more broadly distributed across the CD8_CX3CR1, CD8_GZMK, and CD8_HSPA1A subsets, indicating active T-cell priming and diverse antigenic experience within the lymphoid compartment (**Figure 3E**). In contrast, the top clones in the PB were largely confined to the CD8_CX3CR1 subset, reflecting a circulating cytotoxic effector population that may play a role in immune surveillance and systemic immune responses (**Figure 3F**).

To further characterize T-cell behaviour, we applied the STARTRAC framework to quantify clonal expansion, migration, and state transition indices across CD8⁺ T-cell subsets (**Figure 3G**). The CD8_GZMK and CD8_CX3CR1 subsets exhibited the highest expansion scores, reflecting their strong clonal proliferation in the tissue microenvironment. Interestingly, CD8_HSPA1A cells presented the highest migration and transition indices, suggesting that these LN-resident cells may possess migratory and plastic phenotypes, potentially leading to the generation of more inflammation-prone subsets, such as CD8_GZMK cells. To explore the developmental relationships among CD8⁺ T-cell populations, we performed a PAGA trajectory analysis, which revealed a close connection between the CD8_HSPA1A and CD8_GZMK subsets (**Figure 3H**). Notably, the PAGA graph also indicated connectivity between CD8_GZMK and other CD8 states, including CD8_CCR7 and CD8_CXCR3, suggesting that CD8_GZMK may be reachable through multiple transcriptional routes. This relationship was further supported by the results of the clonal similarity analysis, in which CD8_HSPA1A and CD8_GZMK displayed the highest degree of clonal similarity among all the subsets (**Figure 3I**), supporting CD8_HSPA1A as a major precursor-like state linked to the lung CD8_GZMK population. Multiplex immunofluorescence staining further confirmed a marked increase in CD8⁺HSP70⁺ cells within the lymph nodes of IPF patients compared with those of normal controls, indicating an enhanced stress response and immune activation in the fibrotic microenvironment (**Figure 3J**). These findings suggest that CD8_HSPA1A cells in the lymph nodes may act as transitional intermediates capable of migrating into the fibrotic lung and differentiating into proinflammatory CD8_GZMK cells.

Following our investigation into CD8⁺ T-cell heterogeneity, we further examined the distribution and functional characteristics of CD4⁺ T cells in IPF. The CD4_CCR7 subset, which is characterized by the expression of classical naïve T-cell markers, was predominantly enriched in the LN and PB. The CD4_ANXA2 subset, characterized by elevated expression of *ANXA2*, *ANXA1*, and *ICAM2*, corresponded to central memory-like T cells and was broadly distributed across PB, LN, and pulmonary tissue (PT). In contrast, CD4_CD69 cells expressing the tissue-resident marker *CD69*, CD4_FOXP3 cells expressing the regulatory marker *FOXP3*, and CD4_CXCR5 cells expressing T follicular helper markers (*CXCR5*, *BCL6*, and *ICA1*) were primarily localized to the LN and PT, with the greatest enrichment in the LN (**Supplementary Figure 2A and 2G**).

### CD8_GZMK–Fibroblast Interactions Drive Fibrogenesis in IPF

We performed unsupervised clustering analysis on 2,659 high-quality fibroblasts and classified them into seven distinct subpopulations through manual annotation combined with established marker genes^28,29^ (**Figure 4A**). These comprised four canonical fibroblast subtypes and three specialized subsets: smooth muscle cells (expressing MYH11, TAGLN, and PLN), pericytes (expressing PDGFRB and COX4I2), and mesothelial cells (expressing KRT18, SLPI, and UPK3B) (**Figure 4B**).

**Figure 4.**
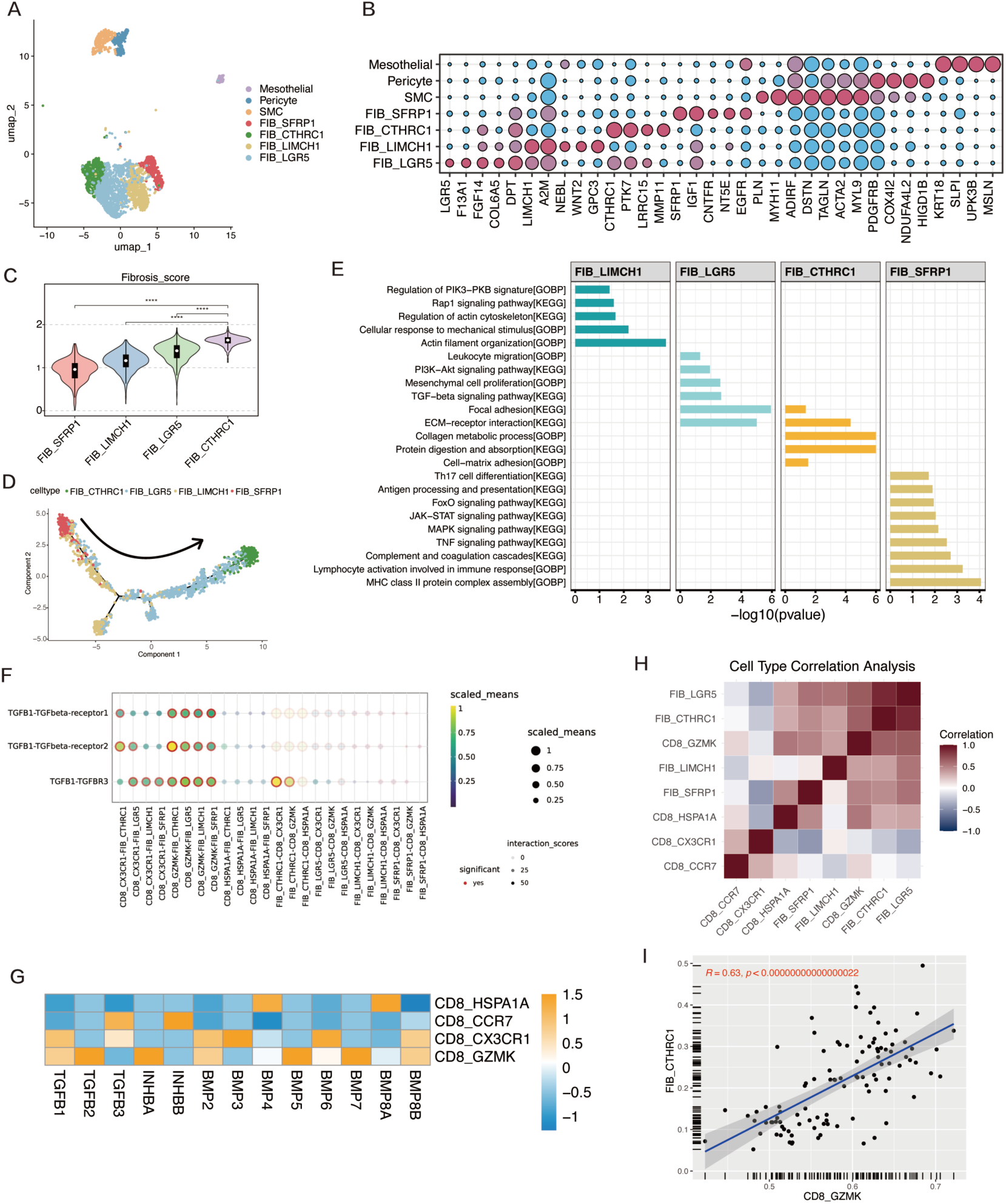
GZMK+CD8+ T cell‒fibroblast crosstalk drives fibrogenesis signalling in IPF. **A.** UMAP plot of fibroblasts from IPF patients, with reclustering into 7 distinct clusters, each represented by a different colour. **B.** Dot plot displaying the expression of marker genes across identified fibroblast clusters. **C.** Violin plot showing fibrosis scores across fibroblast subsets. **D.** Monocle analysis revealing the differentiation trajectories of fibroblast subtypes in IPF. Cells are colour-coded by subtype and include CTHRC1^+^ (green), LGR5^+^ (blue), LIMCH1^+^ (yellow), and SFRP1^+^ (red) fibroblasts. **E.** Bar plot displaying the selected KEGG and GO pathway enrichment results for five fibroblast subsets based on the top 200 genes from each cluster. **F.** Ligand–receptor interaction analysis revealed significant TGFB1–TGFBR1/2/3 signalling interactions between CD8_GZMK subsets and fibroblast populations in IPF lung tissue. **G.** Heatmap showing the expression profiles of TGF-β superfamily ligands (including TGFBs, INHs, and BMPs) across CD8⁺ T-cell subsets. **H.** Heatmap of cell-type correlations based on ssGSVA analysis showing a strong positive correlation between CD8_GZMK cells and the FIB_CTHRC1 fibroblast subset. **I.** Correlation scatter plot of the CD8_GZMK and FIB_CTHRC1 subsets revealed a significant positive correlation, with a Spearman correlation coefficient of 0.63.

To evaluate the fibrotic potential of these fibroblast subsets, we analysed the expression of ECM-related genes and calculated fibrosis scores. The FIB_CTHRC1 population exhibited the highest expression of ACTA2, FN1, and multiple collagens, which is consistent with a robust myofibroblast phenotype (**Supplementary Figure 3A**). Consistent with these findings, FIB_CTHRC1 had the highest fibrosis score among all the fibroblast types, followed by FIB_LGR5 and FIB_LIMCH1, whereas FIB_SFRP1 had the lowest fibrosis score (**Figure 4C, Supplementary Figure 3B**). Monocle-based pseudotime trajectory analysis revealed a progressive trajectory of fibroblast differentiation, with FIB_SFRP1 localized at the origin and FIB_CTHRC1 at the terminal end, supporting a progressive activation process (**Figure 4D, Supplementary Figure 3C**). Pathway enrichment analysis further revealed distinct functional profiles across fibroblast subsets (**Figure 4E**). FIB_LIMCH1 fibroblasts were enriched in pathways related to the mechanical stress response and cytoskeletal organization, such as PI3K–PKB signalling and the regulation of actin dynamics. FIB_LGR5 cells were enriched in mesenchymal proliferation and TGF-β signalling pathways, suggesting a profibrotic profile. The FIB_CTHRC1 subset was associated with extracellular matrix organization and collagen metabolic processes, reinforcing its central function in fibrotic remodelling. Conversely, FIB_SFRP1 fibroblasts were enriched in immune-related pathways, including antigen presentation, JAK-STAT signalling, and complement activation, indicating potential immunoregulatory roles.

To explore the crosstalk between fibroblasts and T cells, we performed ligand–receptor interaction analysis focusing on the TGF-β and CXCL signalling axes. The results revealed that CD8_GZMK cells may interact with the FIB_CTHRC1 and FIB_LGR5 subsets via the TGFβ1–TGFBR1/TGFBR2 axis, suggesting that CD8_GZMK cells promote myofibroblast activation and progression of fibrosis through the TGFβ pathway (**Figure 4F**). In addition, FIB_CTHRC1 cells strongly interacted with CD8_GZMK cells via the CXCL14–CXCR4 and CXCL16–CXCR6 axes, suggesting that fibroblasts may actively recruit CD8_GZMK cells to fibrotic regions through chemokine secretion (**Supplementary Figure 3E**). We next examined the expression of TGF-β superfamily ligands across CD8^+^ T-cell subsets (**Figure 4G**). Notably, the expression of TGFB1 and TGFB2 was elevated in CD8_GZMK cells, suggesting that these cells can directly contribute to TGF-β–mediated fibrotic signalling. Moreover, INHBA expression was specifically and robustly elevated in CD8_GZMK cells, highlighting a possible role for activin A signalling in fibroblast activation.

Finally, ssGSVA of the GSE213001 dataset revealed significantly higher infiltration scores of CD8_GZMK cells in IPF tissues than in normal control tissues (**Supplementary Figure 3D**), indicating disease-associated enrichment of this subset. Correlation analysis of cell type enrichment scores revealed a strong positive correlation between CD8_GZMK cells and the FIB_CTHRC1 fibroblast subset (**Figure 4H**). This relationship was further validated by a significant Spearman correlation (R = 0.63) between the abundance of CD8_GZMK and FIB_CTHRC1 cells, further supporting a potential link between CD8_GZMK cells and fibroblast-mediated fibrogenesis (**Figure 4I**).

### GZMK⁺CD8⁺ T cells are increased in IPF and correlated with fibrosis severity and poor prognosis

To evaluate the clinical significance and fibrotic relevance of CD8⁺GZMK⁺ T cells in IPF, we conducted a clinical validation using peripheral blood and lung tissue samples from IPF patients. Flow cytometry analysis revealed a markedly greater frequency of GZMK⁺CD8⁺ T cells in the peripheral blood of IPF patients than in that of healthy controls (HC) (P = 0.0043; **Figure 5A**). To investigate the prognostic implications of this subset, we stratified patients from the publicly available GSE28042 cohort on the basis of CD8_GZMK signature scores and conducted a Kaplan–Meier survival analysis. Compared with patients with lower scores, patients with higher CD8_GZMK scores had significantly shorter overall survival (P < 0.0001; **Figure 5B**), indicating that elevated numbers of GZMK⁺ CD8⁺ T cells may serve as a predictor of poor clinical outcomes.

**Figure 5.**
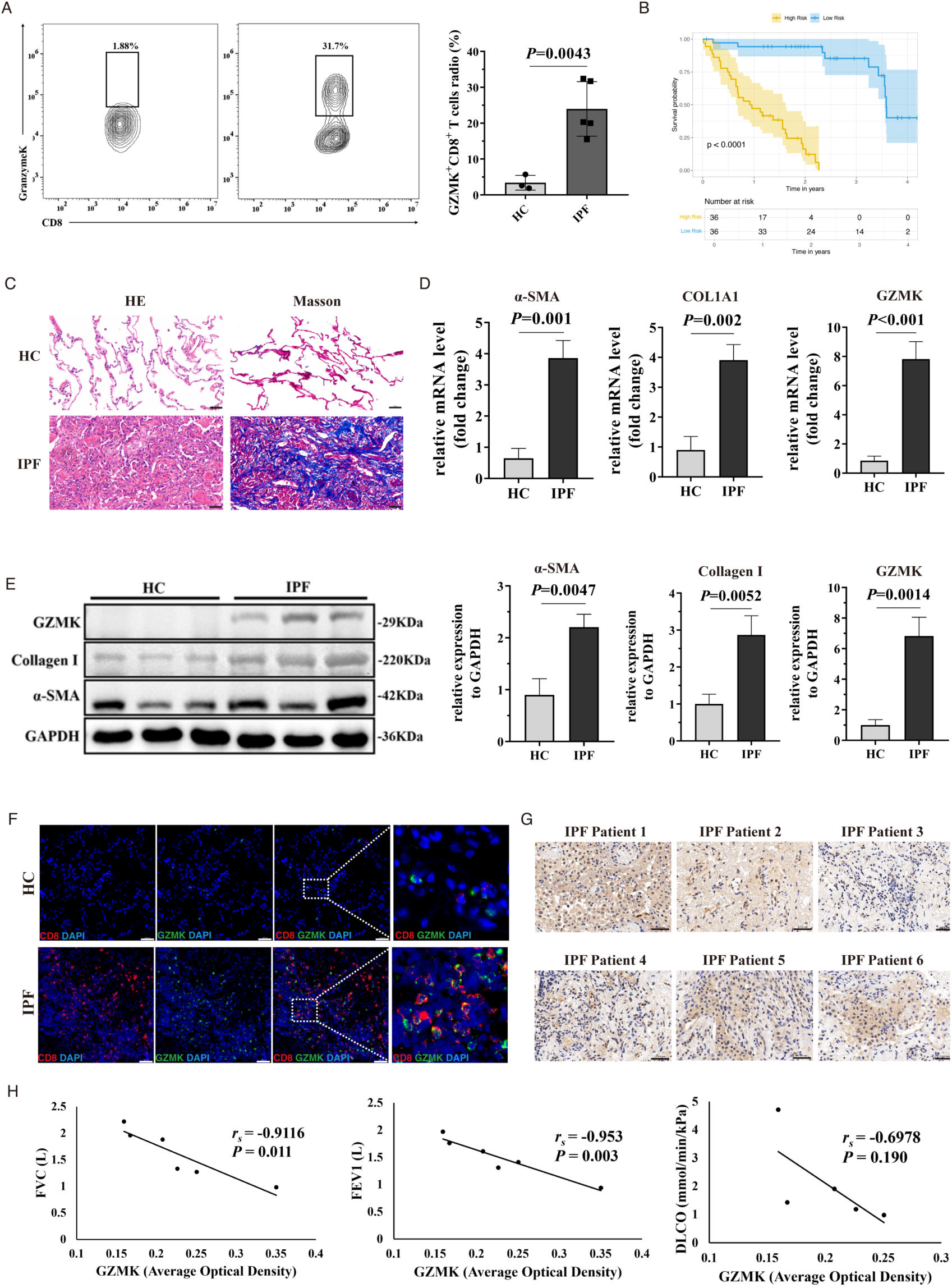
GZMK⁺ CD8⁺ T cells are elevated in IPF and are associated with fibrosis severity and poor prognosis. **A.** Flow cytometry analysis revealed a significantly greater proportion of GZMK⁺CD8⁺ T cells in peripheral blood from IPF patients than in that from healthy controls. **B.** Kaplan‒Meier survival curve analysis of the GSE28042 dataset revealed that IPF patients with higher GZMK⁺CD8⁺ T-cell scores had significantly worse overall survival. **C.** Representative haematoxylin and eosin (H&E) and Masson’s trichrome staining images showing disrupted alveolar architecture and excessive collagen deposition in lung tissue of patients with IPF compared with normal lung tissue. scale bar, 50 μm. **D.** qPCR analysis showing significantly increased mRNA expression of COL1A1, α-SMA, and GZMK in lung tissues from idiopathic pulmonary fibrosis (IPF) patients compared with those from healthy controls (HC). **E.** Western blot analysis and quantification revealed markedly elevated protein levels of collagen I, α-SMA, and GZMK in lung tissues of patients with IPF compared with those in HCs, with GAPDH used as a loading control. **F.** Multiplex immunofluorescence staining showing enrichment of CD8⁺GZMK⁺ double-positive cells (red and green) in IPF lung tissue, but rarely in normal lung tissue. DAPI (blue) was used to stain the nuclei. scale bar, 50 μm. n = 3 individual patient samples were examined independently. **G.** Immunohistochemical staining of **GZMK** in lung tissue sections from six independent IPF patients. Brown signals indicate positive staining for GZMK, with variable expression levels observed across individual patients. Nuclei were counterstained with haematoxylin (blue). Scale bar, 50 μm. **H.** Analysis of the correlation between GZMK expression and pulmonary function parameters in IPF patients. Scatter plots show negative correlations between the average optical density of GZMK staining and forced vital capacity (FVC, left), forced expiratory volume in 1 second (FEV1, middle), and diffusing capacity of the lungs for carbon monoxide (DLCO, right). Spearman’s correlation coefficients (r_s_) and corresponding p values are indicated in each panel. Data are presented as mean ± standard error of the mean (SEM). Statistical significance was determined using two-tailed Student’s t-tests.

Histopathological assessment using haematoxylin and eosin (H&E) and Masson’s trichrome staining of lung sections from IPF patients further revealed severe disruption of alveolar architecture and extensive collagen deposition in IPF lungs, consistent with progressive fibrotic remodelling (**Figure 5C**). Both qPCR and Western blotting revealed significantly elevated expression of fibrosis-related markers (COL1A1 and α-SMA) and GZMK in IPF lung tissue samples compared with control tissue samples. Multiplex immunofluorescence staining revealed marked accumulation of GZMK⁺CD8⁺ T cells in fibrotic lung tissue but not in normal lung tissue (**Figure 5F**), suggesting the selective enrichment and potential activation of this subset within the fibrotic microenvironment.

To further elucidate the functional significance of GZMK expression in relation to disease severity, we analysed the correlation between GZMK transcript levels and clinical pulmonary function parameters, including forced vital capacity (FVC), forced expiratory volume in 1 second (FEV1), and DLCO. The expression of GZMK was significantly negatively correlated with the FVC and FEV1 in IPF patients (**Figure 5G and 5H**), reinforcing its potential role in the pathogenesis of fibrosis and highlighting its utility as a prognostic biomarker.

### GZMK overexpression in CD8⁺ T cells promotes fibroblast activation and proliferation

To determine the direct functional impact of GZMK expression in CD8⁺ T cells on fibroblast behaviour, we established an in vitro coculture model using human foetal lung fibroblasts (HFL1) and human peripheral CD8⁺ T cells genetically modified to overexpress (GZMK-OE) or knockdown (GZMK-shRNA) GZMK (**Figure 6A**). Efficient GZMK modulation in CD8⁺ T cells was confirmed at both the mRNA and protein levels by qPCR and Western blotting, respectively (**Figure 6B and 6C**).

**Figure 6.**
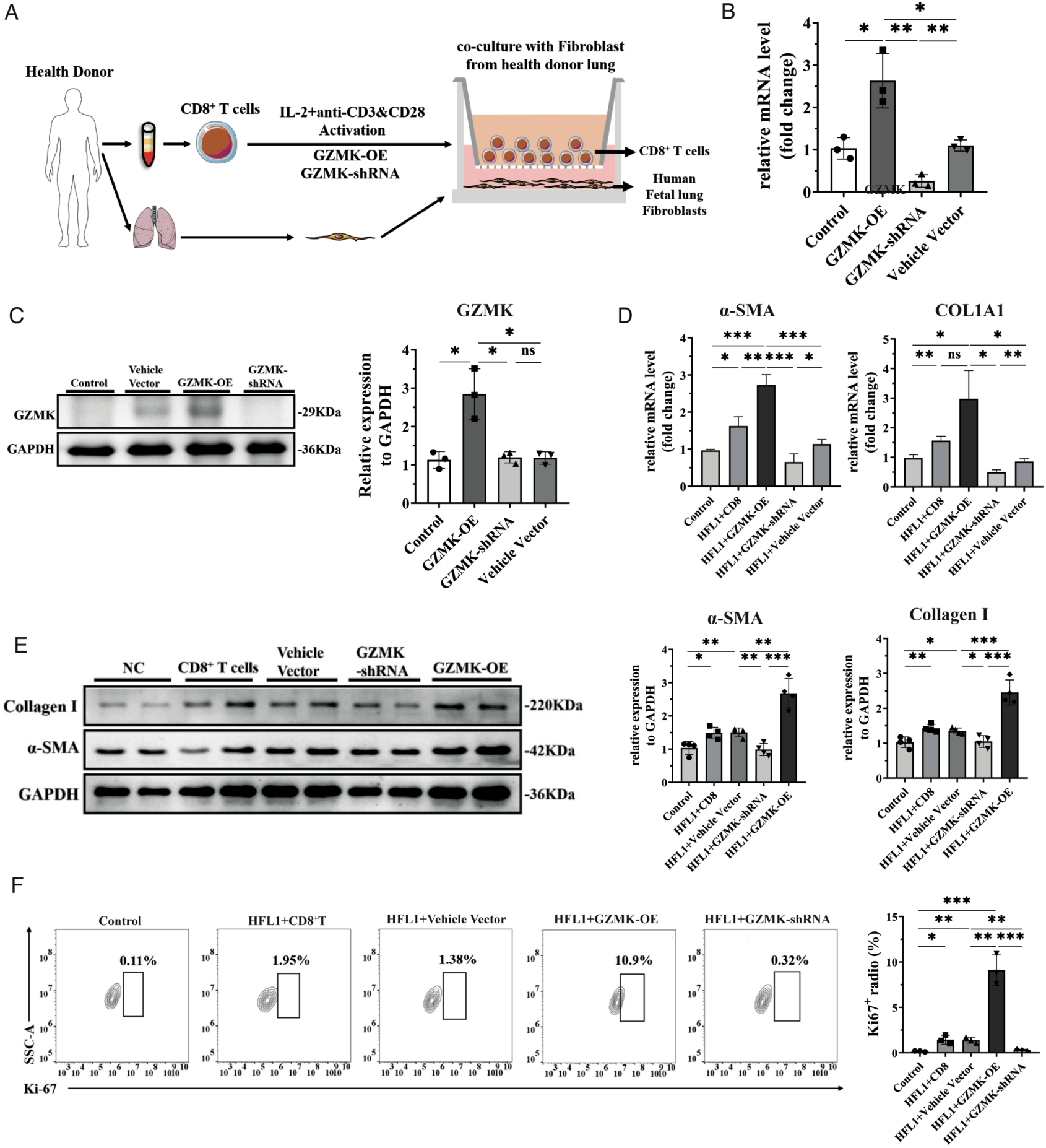
GZMK^+^CD8^+^ T cells directly induce fibroblast-to-myofibroblast transition in human lung fibroblasts. **A.** Experimental workflow: Human foetal lung fibroblasts (HFL1) were cocultured with human peripheral CD8⁺ T cells that had been activated and genetically modified to overexpress (OE) or knockdown (shRNA) GZMK to assess functional effects. **B‒C.** qPCR analysis (B) and Western blotting (C) confirming the successful overexpression and knockdown of GZMK in human CD8⁺ T cells after transduction. **D.** qPCR analysis of fibrosis-related markers (COL1A1 and α-SMA) in fibroblasts cocultured with GZMK-modified CD8⁺ T cells. The overexpression of GZMK promotes fibrotic gene expression, whereas its knockdown suppresses it. **E.** Western blot analysis of collagen I and α-SMA expression in fibroblasts cocultured with different types of CD8⁺ T cells. **F.** Quantification of Ki67⁺ fibroblasts after coculture with different types of CD8⁺ T cells. GZMK overexpression enhances fibroblast proliferation, whereas its knockdown reduces it. Data are presented as mean ± standard error of the mean (SEM). Statistical significance was determined using a one-way ANOVA with multiple comparisons. *P<0.05, **P<0.01, ***P<0.001, ****P<0.0001, ns, not significant.

Coculture of fibroblasts with GZMK-OE CD8⁺ T cells markedly upregulated the expression of the fibrotic genes COL1A1 and α-SMA, whereas GZMK knockdown significantly suppressed the expression of these genes (**Figure 6D**). Consistently, Western blot analysis revealed increased protein levels of collagen I and α-SMA in fibroblasts exposed to GZMK-OE T cells, with corresponding decreases observed following GZMK knockdown (**Figure 6E**). Furthermore, flow cytometric analysis of the expression of Ki-67, a marker of cellular proliferation, revealed that GZMK-OE CD8⁺ T cells significantly enhanced fibroblast proliferation, whereas GZMK-shRNA CD8⁺ T cells inhibited proliferation (**Figure 6F**).

Collectively, these findings provide direct functional evidence that GZMK overexpressing CD8⁺ T cells can drive fibroblast-to-myofibroblast transition and promote fibroblast proliferation, implicating GZMK as a key effector in profibrotic immune–stromal interactions.

### Inhibition of GZMK Suppressed Fibroblast-to-Myofibroblast Transition and Attenuated Pulmonary Fibrosis

To further validate the pathogenic role of GZMK in fibroblast activation and pulmonary fibrosis, we investigated the therapeutic effect of bosutinib, a Src family kinase inhibitor known to suppress GZMK activity. A coculture system was established in which HFL1 cells were exposed to peripheral blood mononuclear cells (PBMCs) from IPF patients to simulate the immune-fibrotic microenvironment (**Figure 7A**). Quantitative PCR analysis and Western blotting demonstrated that coculture with IPF-derived PBMCs significantly upregulated the expression of the fibrotic markers α-SMA and COL1A1 in HFL1 cells, whereas treatment with bosutinib (10 µM) markedly reduced their expression (**Figure 7B**, **Supplementary Figure 4C**). These findings indicate that GZMK inhibition can mitigate immune cell–induced fibroblast-to-myofibroblast transition. Flow cytometry analysis revealed that the percentage of Ki-67⁺ proliferating fibroblasts was significantly elevated upon exposure to IPF-derived PBMCs, whereas bosutinib treatment led to a substantial decrease in the number of Ki-67⁺ cells, indicating that GZMK activity is associated with enhanced fibroblast proliferation (**Figure 7C, Supplementary Figure 4D**).

**Figure 7.**
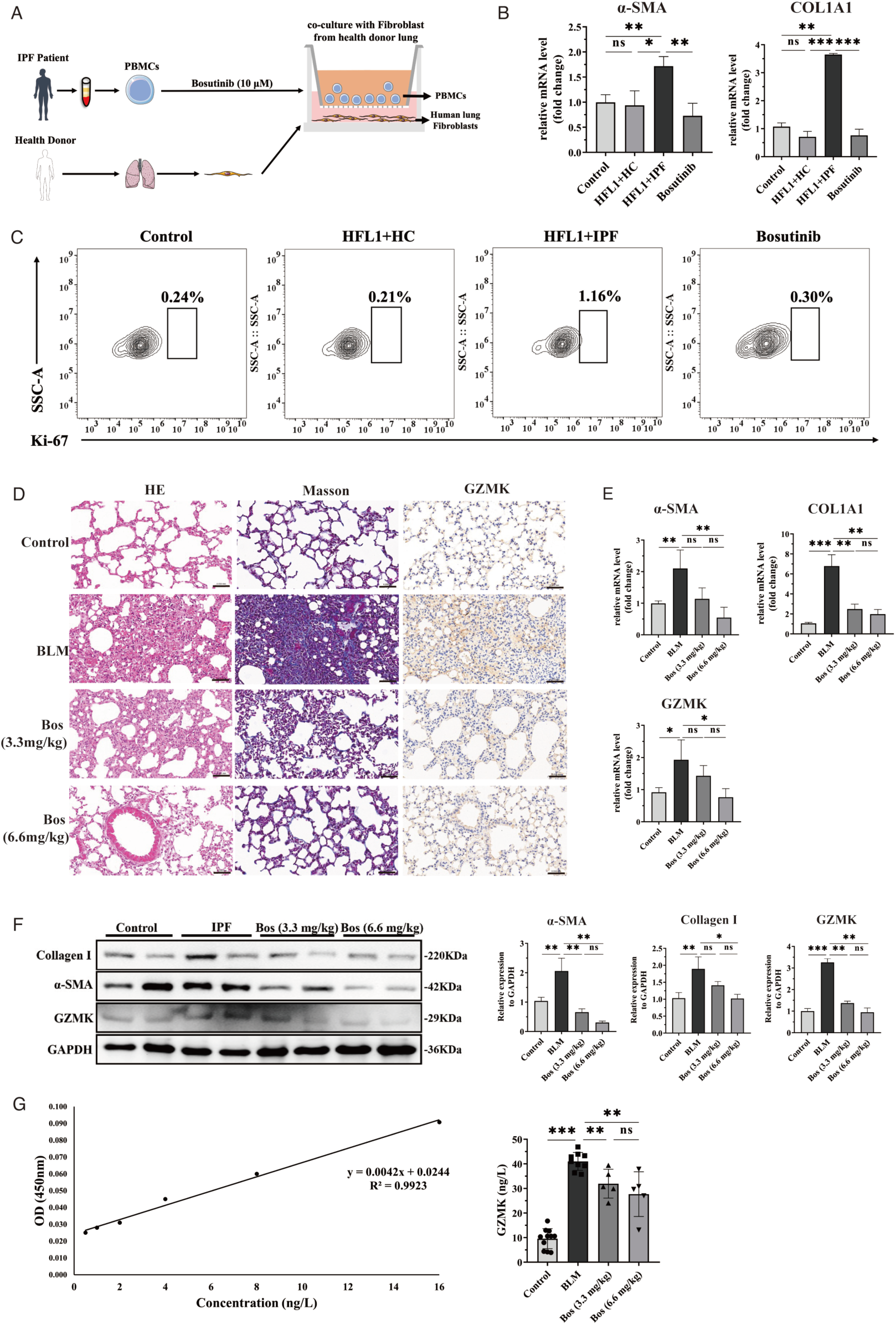
GZMK inhibition reduces fibroblast-to-myofibroblast transition and alleviates pulmonary fibrosis. **A.** Schematic diagram of GZMK inhibition in IPF patient-derived PBMC and HFL1 cell coculture chambers. **B.** qPCR analysis showing the expression levels of the fibrotic markers COL1A1 and α-SMA in fibroblasts cocultured with IPF-derived PBMCs with or without treatment with the GZMK inhibitor bosutinib. **C.** Quantification of Ki67-positive fibroblasts after coculture with PBMCs. Treatment with bosutinib significantly decreased fibroblast proliferation. **D.** BLM-induced lung fibrosis mice were subsequently treated with bosutinib every 3 days (3.3 or 6.6 mg/kg, intraperitoneally) to evaluate its in vivo antifibrotic effects. Representative images of HE, Masson’s trichrome, and immunohistochemical staining for GZMK in lung tissues from different groups. scale bar, 50 μm. **E.** qPCR analysis further demonstrated that bosutinib significantly decreased the expression of α-SMA, COL1A1, and GZMK. **F.** Western blot analysis confirmed that bosutinib effectively decreased the protein levels of collagen I, α-SMA, and GZMK in lung tissue homogenates, which is consistent with its antifibrotic effects. **G.** Serum GZMK protein levels in the indicated treatment groups were quantified by ELISA. Data are presented as mean ± standard error of the mean (SEM). Statistical significance was determined using a one-way ANOVA with multiple comparisons. *P<0.05, **P<0.01, ***P<0.001, ****P<0.0001, ns, not significant. Bos, Bosutinib. BLM, Bleomycin.

In a BLM-induced murine model of pulmonary fibrosis, histological evaluation using H&E and Masson’s trichrome staining revealed severe alveolar destruction and collagen deposition in BLM-treated lungs, which were markedly alleviated by bosutinib in a dose-dependent manner (3.3 mg/kg and 6.6 mg/kg, respectively) (**Figure 7D**). Immunohistochemical staining for GZMK revealed prominent expression in fibrotic lungs, which was markedly reduced following bosutinib treatment. qPCR analysis revealed that bosutinib at 6.6 mg/kg significantly reduced α-SMA and GZMK mRNA levels compared with BLM alone, whereas COL1A1 expression was decreased at both 3.3 and 6.6 mg/kg (**Figure 7E**). (**Figure 7E**). Western blotting demonstrated that collagen I, α-SMA and GZMK protein levels were lower in lungs from mice treated with bosutinib at 6.6 mg/kg than in BLM-treated mice, while only α-SMA, and GZMK were reduced at 3.3 mg/kg (**Figure 7F**). (**Figure 7F**). Densitometric quantification revealed that bosutinib at 6.6 mg/kg exerted a more pronounced suppressive effect on the expression of fibrotic markers, indicating dose-dependent pharmacological efficacy. To directly quantify GZMK protein levels in lung homogenates, ELISA was performed. A standard curve confirmed the assay linearity (**Figure 7G**), and analysis of tissue samples revealed significantly increased GZMK levels in the BLM group, which were markedly reduced by bosutinib in a dose-dependent fashion (**Figure 7G**).

Collectively, these results demonstrate that inhibition of GZMK by bosutinib effectively suppresses fibroblast activation and proliferation, thereby attenuating the progression of pulmonary fibrosis. These findings further support GZMK as a key mediator of disease progression.

### Pharmacologic Blockade of TGFβR1/ALK4 Mitigates Fibrogenic Responses in Lung Fibroblasts and In Vivo Models

To investigate potential mediators downstream of GZMK⁺CD8⁺ T cells in the regulation of fibrotic responses, we first examined the expression of TGF-β pathway components in human lung tissues. Quantitative PCR analysis revealed significant upregulation of the expression of ALK4, a type I receptor in the TGF-β superfamily, in lungs of IPF patients compared with that in healthy control lungs, while the expression of TGFβR1 remained unchanged (**Supplementary Figure 4A**). We next assessed whether GZMK-modified CD8⁺ T cells could influence receptor expression in fibroblasts in vitro. Fibroblasts cocultured with GZMK-OE CD8⁺ T cells exhibited markedly increased mRNA levels of both TGFβR1 and ALK4, whereas GZMK knockdown suppressed their expression (**Supplementary Figure 4B**), suggesting that GZMK⁺CD8⁺ T cells may promote fibroblast activation in part through upregulation of the expression of these TGF-β receptors.

To investigate whether pharmacologic inhibition of TGFβR1/ALK4 could mitigate fibroblast activation, we cocultured human lung fibroblasts with PBMCs derived from IPF patients and treated the cultures with TEW (vactosertib, TEW-7197), a selective TGFβR1/ALK4 inhibitor. qPCR and western blot revealed that, compared with no treatment, TEW treatment significantly reduced the expression of key fibrotic markers, including COL1A1 and α-SMA, in fibroblasts (**Figure 8A**, **Supplementary Figure 4C**). Consistently, flow cytometric analysis revealed a marked reduction in the proportion of Ki-67⁺ proliferating fibroblasts upon TEW treatment, suggesting suppressed fibroblast proliferation (**Figure 8B**, **Supplementary Figure 4D**).

**Figure 8.**
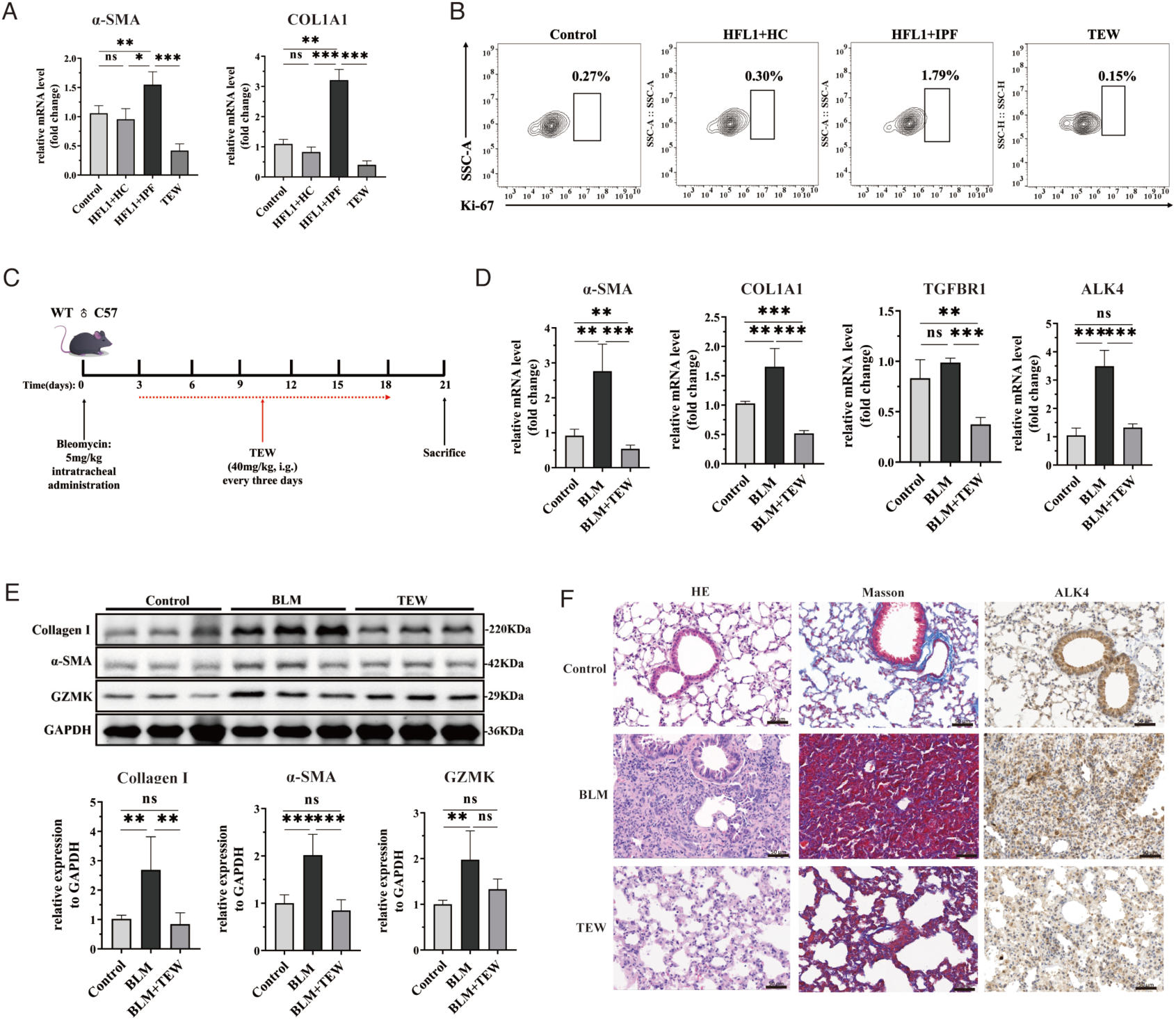
The TGFβR1/ALK4 inhibitor TEW prevents fibroblast activation and fibrotic progression *in vitro* and *in vivo*. **A.** qPCR analysis of fibrotic marker genes (COL1A1, α-SMA) in normal lung fibroblasts co-cultured with PBMCs isolated from patients with idiopathic pulmonary fibrosis (IPF), with or without TEW-7197 treatment. **B.** Quantification of fibroblast proliferation (assessed by Ki67-positive staining) following co-culture with IPF-derived PBMCs, with or without TEW treatment. **C.** Schematic of the in vivo experimental timeline. Pulmonary fibrosis was induced in C57BL/6 mice via intranasal instillation of bleomycin (5 mg/kg). Mice were treated with TEW (40 mg/kg, p.o.) or vehicle every three days thereafter. **D.** qPCR analysis of fibrotic and target genes (α-SMA, COL1A1, TGFΒR1, ALK4) in lung tissue from bleomycin-challenged mice treated with TEW or vehicle control. **E.** Western blot analysis (left) and corresponding densitometric quantification (right) of Collagen I and α-SMA protein levels in mouse lung tissue. GZMK served as a loading control. **F.** Representative photomicrographs of lung sections stained with Hematoxylin and Eosin (H&E; top), Masson’s Trichrome (middle; collagen in blue), and for ALK4 by immunohistochemistry (IHC; bottom). scale bar, 50 μm. Data are presented as mean ± standard error of the mean (SEM). Statistical significance was determined using a one-way ANOVA with multiple comparisons. *P<0.05, **P<0.01, ***P<0.001, ****P<0.0001, ns, not significant. TEW, TEW-7197.

To assess the antifibrotic efficacy of TEW in vivo, we used a BLM-induced pulmonary fibrosis model in C57BL/6 mice and orally administered TEW every three days (**Figure 8C**). qPCR analysis of lung tissues revealed that compared with no treatment, TEW treatment significantly decreased the expression of the fibrotic markers α-SMA and COL1A1, as well as the expression of the TGF-β receptors TGFβR1 and ALK4, although TGFβR1 expression was not significantly elevated by BLM exposure (**Figure 8D**). Western blot analysis further confirmed the decreased protein levels of collagen I and α-SMA in TEW-treated lungs, whereas GZMK expression remained unchanged (**Figure 8E**). Histological assessment via H&E and Masson’s trichrome staining revealed marked attenuation of alveolar disruption and collagen deposition in TEW-treated mice. Additionally, immunohistochemical staining revealed reduced ALK4 expression following TEW administration (**Figure 8F**).

Together, these findings demonstrated that TGFβR1/ALK4 inhibition via TEW effectively restrained fibroblast activation and fibrosis progression both in vitro and in vivo, supporting its therapeutic potential in IPF.

## Discussion

In this study, we performed single-cell transcriptomic profiling and TCR repertoire analysis on paired samples from the lung parenchyma, mediastinal lymph nodes, and peripheral blood of patients with IPF and identified a previously unrecognized GZMK⁺CD8⁺ T-cell subset with a proinflammatory but low-cytotoxicity phenotype. This subset is highly enriched and clonally expanded in fibrotic lungs and closely related to lymph node–resident CD8_HSPA1A cells. Functional assays demonstrated that GZMK overexpressing CD8+ T cells promote fibroblast-to-myofibroblast differentiation and proliferation via the TGF-β1 signalling axis. The inhibition of GZMK expression significantly attenuated fibroblast activation and collagen deposition. Furthermore, the selective blockade of TGFβR1/ALK4 using TEW suppressed fibroblast activation in vitro and mitigated disease progression in a BLM-induced mouse model of pulmonary fibrosis, indicating the therapeutic potential of targeting this pathway.

The pathogenesis of IPF remains incompletely understood. Emerging evidence supports the notion that chronic inflammation underlies fibrotic remodelling, where persistent inflammatory insults cause recurrent alveolar injury, leading to fibroblast proliferation, differentiation, and ECM deposition^30,31^. We identified a critical Tem subset called CD8_GZMK, which is significantly enriched in IPF lung tissue. This population is transcriptionally regulated by *JUN, JUND*, and *EGR1* and is characterized by low cytotoxicity but high proinflammatory cytokine secretion. This phenotype mirrors GZMK⁺CD8⁺ T cells found in chronic viral infections^32^, senescence-associated secretory phenotype (SASP)^33^ and tumour microenvironments^14^, where they have been implicated in sustaining inflammation. These findings collectively point to GZMK⁺ T cells as a conserved, tissue-associated T-cell subset across multiple chronic inflammatory diseases. Our study is the first to demonstrate their direct involvement in profibrotic immune–stromal interactions in IPF.

Additionally, we identified a lymph node-specific CD8⁺ T-cell subset, CD8_HSPA1A, which is highly enriched in lymph nodes. TCR repertoire analysis revealed that this subset has the greatest migratory and differentiation potential and shares strong clonal similarity and developmental trajectory with CD8_GZMK cells. Notably, the lymph node microenvironment, which is enriched in heat shock proteins (HSPs), may facilitate T-cell activation and migration^34,35^. These observations suggest that CD8_HSPA1A cells may undergo initial activation in lymph nodes, migrate to the lung, and differentiate into CD8_GZMK cells, contributing to tissue injury and inflammation. This potential migratory differentiation pathway may also help explain the frequently observed MLNE in IPF and underscores the critical role of lymph nodes in IPF pathogenesis.

In line with previous reports^28^, our fibroblast analysis revealed highly invasive CTHRC1⁺ fibroblasts (coexpressing ACTA2) and early SFRP1⁺ damage-activated fibroblasts. Bidirectional interactions between GZMK⁺CD8⁺ T cells and fibroblasts were observed. Fibroblasts secrete chemokines (CXCL13 and CXCL16) that recruit and retain CD8_GZMK cells within fibrotic lesions, which is consistent with the localized immune activation observed in chronic inflammation. In turn, GZMK⁺ T cells express high levels of TGF-β1 and activin A (INHBA), enhancing fibrotic gene expression and TGF-β receptor signalling in fibroblasts, thereby accelerating myofibroblast transition via paracrine mechanisms. This fibrogenic function complements previous findings regarding the role of CD4⁺ T cells and Th2 cells in fibrosis^36,37^, we provides the first systematic evidence of a noncanonical fibrotic role for GZMK^+^CD8⁺ T cells mediated by TGFβ signalling.

Our in vitro coculture model confirmed the pathogenic role of GZMK⁺CD8⁺ T cells in IPF by demonstrating their ability to directly promote fibroblast activation and proliferation and to drive their transition into myofibroblasts. The use of bosutinib, a Src kinase inhibitor known to suppress GZMK activity, further validated the functional contribution of GZMK to IPF pathogenesis^38^. While traditionally associated with cytotoxic activity, GZMK has been increasingly recognized for its nonlytic functions in inducing proinflammatory responses and transcriptional reprogramming in target cells^13^. Our findings not only elucidate the functional landscape of CD8⁺ T cells in tissue remodelling but also highlight GZMK as a key immune mediator of fibrosis, in line with recent studies on the noncytolytic roles of granzymes in chronic inflammation and tissue damage.

Currently, no TGF-β receptor-specific inhibitors have been approved for IPF treatment. However, emerging studies have identified ALK4 (activin receptor-like kinase 4), a type I receptor of the TGF-β superfamily, as a promising therapeutic target. Although ALK4 shares downstream signalling pathways with the TGF-β receptor, particularly through Smad2/3 activation, it has distinct biological functions. ALK4 primarily mediates activin A signalling and has recently been implicated in immune regulation^39^. Notably, ALK4 expression in CD4⁺ T cells has been shown to promote the differentiation of pathogenic Th17 cells, a subset associated with autoimmune disease^40,41^. Whether ALK4 activation in CD8⁺GZMK⁺ T cells contributes similarly to fibrosis remains to be determined. In our in vivo studies, pharmacological inhibition of ALK4 effectively attenuated lung fibrosis, suggesting that it plays a direct role in disease progression. Moreover, ALK4 inhibition in cocultures of HLF1 cells with IPF patient-derived PBMCs significantly suppressed fibrotic gene expression. These findings support the use of selective ALK4 blockade as a potentially targeted antifibrotic strategy in IPF.

Despite these novel insights, this study has several limitations. First, our single-cell cohort included only four male IPF patients undergoing lung transplantation, with heterogeneous smoking histories and treatment regimens; therefore, it does not capture the full clinical and molecular heterogeneity of IPF or allow robust stratification by sex, smoking burden or treatment status. Although this design enabled high-quality matched lung, lymph node and blood profiling, future studies in larger and more diverse cohorts, including female patients and clinically defined subgroups, will be required to confirm the generalizability of our findings. Second, our coculture experiments used CD8⁺ T cells engineered to overexpress or knock down GZMK, showing that GZMK expression in CD8⁺ T cells is sufficient to enhance fibroblast activation in vitro but not proving that endogenous GZMK⁺CD8⁺ T cells, or CD8⁺ T cells themselves, are required for fibroblast activation in vivo. Third, IPF patients differed in background clinical status and medication exposure, including antifibrotic therapy, which may act as potential confounders. Finally, while we delineate a profibrotic role for GZMK⁺CD8⁺ T cells, the mechanisms underlying their activation, antigen specificity and upstream inducers remain to be explored, and future studies should investigate these cells across different pathological subtypes of IPF and assess the safety and specificity of targeting this pathway.

In conclusion, our integrated single-cell and functional analyses revealed that GZMK⁺CD8⁺ T cells promote fibroblast activation and lung fibrosis through the TGFβR1/ALK4 axis, revealing a critical immune–stromal interaction loop involved in IPF progression. These findings offer a novel conceptual framework and therapeutic rationale for targeting the TGFβR1/ALK4 signalling pathway in antifibrotic interventions.

## Data availability

The scRNA-seq and scTCR-seq data supporting the findings of this study have been deposited at GSA-Human (HRA013103) under accession code PRJCA032809. Previously published microarray data analyzed together are available under accession codes GSE28042 and GSE213001. All other supporting data of this study are available from the corresponding author on reasonable request.

## Acknowledgements

We thank all patients and their families for participating in this study.

## Author contributions

J.Y.C., J.Y. and M.H. conceived the study and designed the experiments. B.Q.Y. and Q.X.L. performed bioinformatic data analysis. B.Q.Y., S.L.J, G.Y.X and J.C performed the experiments. B.Q.Y., S.L.J., T.Y., G.G.Z, Q.F.Y. and H.J.Y. interpreted the results. B.H., J.Z., X.C.Y., P.Y., J.H. and J.Y.C. conducted surgery and collected samples from patients. B.Q.Y., S.L.J, and Q.X.L. write the manuscript. J.Y., M.H., G.F. and J.Y.C. reviewed, and revised the manuscript. J.Y., M.H. and J.Y.C. were responsible for material support and supervision. All authors approved the final manuscript.

## Methods

### Ethics approval

This study was performed in accordance with the Declaration of Helsinki and approved by the Ethics Committee of The Second Affiliated Hospital Zhejiang University School of Medicine (ID: 2024-1416). All participants provided the written informed consent before inclusion. All animal study protocols were reviewed and approved by Animal Care & Use Committee of The Second Affiliated Hospital Zhejiang University School of Medicine (ID: 2024-324)

### Study Population and Specimen Collection

Four male patients pathologically diagnosed with IPF who underwent lung transplantation at The Second Affiliated Hospital Zhejiang University School of Medicine were enrolled in this study. Inclusion criteria were: (i) a multidisciplinary and histopathological diagnosis of IPF according to international ATS/ERS/JRS/ALAT guidelines; (ii) eligibility for lung transplantation; and (iii) age ≥ 18 years with the ability to provide written informed consent. Major exclusion criteria were: (i) evidence of connective tissue disease–associated ILD or other defined forms of interstitial lung disease; (ii) active pulmonary infection or malignancy at the time of transplantation; and (iii) prior lung transplantation. For each patient, peripheral blood was collected immediately before surgery, and matched fresh lung tissue and subcarinal mediastinal lymph nodes were obtained intraoperatively immediately after surgical resection. All specimens were promptly processed for single-cell analysis. Clinical characteristics and concomitant medications of the study participants are detailed in Supplementary Table 1. For validation studies using immunofluorescence staining, we conducted the staining procedure on IPF lung and lymph node specimens from the surgical cohort, along with control tissue from unused donor lungs. An additional cohort of six patients with diagnosed IPF was enrolled for Spearman correlation analysis between GZMK expression levels in lung tissues and pulmonary function parameters.

### Sample processing and scRNA sequencing

Fresh tissue specimens (lung and lymph nodes) were processed immediately after collection. Samples were washed in phosphate-buffered saline (PBS; Sangon) to remove blood, adipose tissue, and surface contaminants, then minced into approximately 0.5 mm^2^ fragments in RPMI-1640 medium (GNM11835-500, GENOM) supplemented with 5% PBS. Tissue digestion was performed at 37°C for 20 minutes using an enzymatic cocktail containing 0.35% collagenase type IV (Sigma, C5138-1G), 2 mg/mL papain (Sangon, A003124-0100), and 120 Units/mL DNase I (Sangon, A510099-0001). The resulting cell suspension was filtered through 70 μm strainers and centrifuged at 300g for 5 minutes at 4°C. Erythrocytes were lysed using erythrocyte lysis solution (130-094-183, Miltenyi) for 5 minutes at room temperature. Dead cells were removed using Dead Cell Removal MicroBeads (130-090-101, Miltenyi) according to the manufacturer’s protocol. The final cell pellet was resuspended in PBS containing 0.04% BSA (Sigma, A1933-100G). Peripheral blood samples, peripheral blood mononuclear cells (PBMCs) were isolated within 2 hours of collection using Histopaque-1077 density gradient centrifugation (Sigma) following manufacturer’s guidelines. Isolated PBMCs were resuspended in RPMI-1640 medium supplemented with 5% Fetal Bovine Serum (FBS) (Sangon, E600001-0500). For all samples, single-cell suspensions were adjusted to a final concentration of 700-1,200 cells/μL. Single-cell gene expression profiling and immune repertoire analysis were performed using the Chromium NextGEM Single Cell 5’ Reagent Kits v2(10x Genomics) according to standard protocols. Libraries were sequenced on an Illumina NovaSeq 6000 platform to achieve a minimum depth of 20,000 reads per cell.

### Data processing, quality control, normalization and clustering

Raw sequencing data were processed using the 10x Genomics Cell Ranger pipeline (v7.2.0). Reads were aligned to the human reference genome (GRCh38) and quantified to generate gene-cell count matrices. We implemented stringent quality control filters to remove low-quality cells. Specifically, cells were excluded if they expressed fewer than 200 unique genes or if their mitochondrial content exceeded 25% of total unique molecular identifier (UMI) counts. Potential cell doublets were computationally identified and removed using DoubleFinder (v2.0.3)^42^. Gene expression data were processed using Seurat (v5.1.0). Raw counts were normalized using the “NormalizeData” function, and the top 2,000 highly variable genes were identified for downstream analyses. Data were scaled using the “ScaleData” function, followed by principal component analysis (PCA) using 30 components. To correct for batch effects across tissue types (lung tissue, lymph nodes, and peripheral blood), we employed Harmony (v1.2.0) for data integration^43^. Unsupervised graph-based clustering was performed using Seurat’s “FindClusters” function across multiple resolution parameters (0.5-1.2). Cluster results were visualized using Uniform Manifold Approximation and Projection (UMAP). Differential gene expression analysis between clusters was conducted using the Wilcoxon rank sum test (logFC threshold > 0.25, adjusted P < 0.05). Cell type identities were assigned based on established marker genes from the CellMarker database and previous literature. For detailed analysis of specific cell populations, relevant clusters were subset into new Seurat objects using the “Subset” function.

### Tissue distribution of clusters

To characterize the tissue distribution patterns of cellular populations, we quantified both the relative abundance and tissue-specific enrichment of each cell cluster. For relative abundance, we calculated the proportion of each cell type within the total cell population across samples. To determine tissue specificity, we computed an enrichment score using the ratio of observed to expected cell numbers (Ro/e)^44^, following established methods. Expected cell numbers for each cluster-tissue combination were calculated using chi-squared test assumptions. Clusters with Ro/e values greater than 1 were considered enriched in the corresponding tissue type. This approach enabled systematic identification of tissue-preferential cell populations while controlling for differences in sampling depth across tissues.

### TCR analysis

We analyzed T cell receptor (TCR) sequences using Cell Ranger v7.2.0 (10x Genomics) with the GRCh38-alts-ensembl reference genome. Single-cell TCR sequences were matched to corresponding cellular transcriptomes using unique cell barcodes. Of the analyzed cells, 84.9% of T cells (32,125) yielded high-quality receptor sequences. Clonotypes were inferred using a CDR3 amino-acid–based definition, such that cells sharing identical productive CDR3 amino-acid sequences were assigned to the same clonotype. Expanded (clonal) T cells were defined as clonotypes observed in ≥2 cells, whereas singleton clonotypes (observed in only one cell) were classified as non-expanded (non-clonal). Cells lacking productive TCR contigs were excluded from clonotype-based analyses. Clonal overlap between different CD8⁺ T-cell subsets was quantified by computing a Morisita-Horn similarity index (0–1). The similarity matrix was visualized as a heatmap. We employed STARTRAC v0.1.0 to quantify three key metrics of T cell dynamics: clonal expansion (STARTRAC-expa), tissue migration (STARTRAC-migr), and state transition (STARTRAC-tran)^44^.

### Cell developmental trajectory analysis

To elucidate developmental relationships between cell populations, we employed three complementary computational approaches: Monocle2^45^, and partition-based graph abstraction (PAGA)^46^. Cell ordering along pseudo-temporal trajectories was inferred using Monocle2 (v2.30.1), with cells arranged in a branched structure based on transcriptional similarities. PAGA analysis was implemented through the Scanpy package to map the abstract graph topology of cellular transitions while preserving the global structure of the data. Statistical significance of edge connections between subpopulation nodes was assessed using unpaired two-sided Student’s t-tests across all edges.

### Functional pathway analysis

To characterize cell type-specific functional states, we curated published gene signatures associated with key biological processes. Pathway enrichment scores were calculated for individual cells using the AddModuleScore function in Seurat (v5.1.0) and visualized using UMAP dimensional reduction.

### Differential expression and pathway analysis

To identify cell type-specific markers, we performed differential expression analysis using Seurat’s “FindAllMarkers” function with Wilcoxon rank-sum tests. Genes with adjusted *P* < 0.05 were considered significant markers for each cluster. For functional annotation, we conducted Gene Ontology (GO) and Kyoto Encyclopedia of Genes and Genomes (KEGG) pathway enrichment analyses using clusterProfiler (v4.12.6)^47^.

### Cell-cell interaction analysis

To infer intercellular communication networks, cell-cell communication analysis was performed using CellPhoneDB (v5.0.1) with the v5.0.0 database. Normalized gene expression matrices and annotated cell identities were used as input. For each pairwise combination of cell types, statistically significant ligand-receptor interactions were inferred based on the expression of ligands in one cell type and corresponding receptors in another. Significance was assessed via permutation testing (n = 1000, *P* < 0.05). The number and strength of predicted interactions were visualized using circle plots. For key signaling pathways, selected ligand-receptor pairs were highlighted to illustrate specific intercellular communication patterns.

### Gene regulatory network analysis

To identify active transcription factors (TFs) and their downstream targets within fibroblast subpopulations, we implemented SCENIC (Single-Cell rEgulatory Network Inference and Clustering) v1.3.1^48^. The analysis pipeline consisted of three main steps: First, we used GRNBoost to construct co-expression networks between TFs and their putative target genes from the raw count matrix. Next, we employed RcisTarget to identify direct regulatory relationships by evaluating the enrichment of TF binding motifs in the regulatory regions of co-expressed genes, resulting in high-confidence regulons. Finally, we quantified regulon activity scores for individual cells using AUCell and visualized the results to reveal cluster-specific regulatory programs. This approach enabled systematic identification of master regulators driving fibroblast heterogeneity in IPF.

### Validation using public RNA sequencing datasets

To validate single-cell findings at the transcriptomic level, we analyzed three independent RNA sequencing datasets (GSE28042 and GSE213001). The GSVA package (v1.52.3) was employed to calculate cellular subtype enrichment profiles across individuals, utilizing a gene signature set comprising the top 50 differentially expressed markers for each defined cell cluster. Survival outcomes were assessed through multivariable Cox proportional hazards regression, with Kaplan-Meier survival curves and time-dependent receiver operating characteristic (ROC) analyses generated using the R survival package (v3.7.0) in GSE28042 dataset. The GSE213001 dataset was used for transcriptomic correlation analysis and to assess the infiltration levels of specific T cell subsets. The Pearson correlation between the abundance of GZMK⁺ T cells and CTHRC1⁺ fibroblasts was calculated, and a cell type–level correlation matrix was generated based on enrichment scores across individuals.

### GZMK recombinant plasmid construction

According to the nucleotide sequence of human GZMK (NM_002104) from NCBI databases. We constructed a recombinant lentivirus plasmid (pcDNA3.1-Cppt-IRES) with full length nucleotide sequence of GZMK (795 bp, from 5’ to 3’). The GZMK full-length plasmids was purchased from Vigene Bioscience. In preparation for lentiviral supernatant, before transduction, HEK293T cells were seeded at 6 × 10^6^ cells per 100 mm dish with 80% cell confluency. The lentiviruses were packaging by co-transfecting HEK293T cells with a lentivirus plasmid containing GZMK gene sequence or GZMK-shRNA plasmid, encoding the VSV-G envelope protein plasmid, and the packaging plasmid psPAX2, using PEI transfection mediator. The fresh medium was added after 8 hours. After 48 h, the lentivirus supernatants were all collected and filtered using a 0.45 μm membrane to remove cell debris. Then the lentiviruses were concentrated by ultracentrifugation (Optima XPN-100, Beckmann) at 827,000 × g for 2 h at 4 °C.

### Isolation and culture of human CD8^+^T cells

Peripheral blood mononuclear cells (PBMCs) were obtained from healthy donors by Ficoll-Hypaque (TBD Science) density-gradient centrifugation. Cell number and viability were assessed by trypan blue exclusion, and only PBMC preparations with viability >90% were used for downstream experiments. CD8⁺ T cells were negatively isolated from PBMCs using the CD8⁺ T-lymphocyte enrichment set DM (BD-IMag™, 557941) according to the manufacturer’s instructions, and then activated with 10 ng/mL recombinant human IL-2 (eBioscience), 1 μg/mL anti-CD3 (eBioscience) and 1 μg/mL anti-CD28 (eBioscience) antibodies for 48 h. After lentiviral transduction, CD8⁺ T cells were cultured and expanded in RPMI 1640 (Gibco) supplemented with 10% FBS (Gibco) and 1% penicillin–streptomycin solution (Yeasen), in the continued presence of 10 ng/mL IL-2, 1 μg/mL anti-CD3 and 1 μg/mL anti-CD28 until use in co-culture assays.

### CD8^+^T cells and human fatal lung fibroblasts co-culture assay

In vitro non-contact co-culture assays were used to study the interaction between CD8⁺ T cells and human fetal lung fibroblasts (HFL1). HFL1 cells were maintained in F-12K Nutrient Mixture (Gibco, 21127022) supplemented with 10% FBS (Gibco) and 1% penicillin–streptomycin and were used at passages 2–5. An amount of 5 × 10⁵ HFL1 cells per well was seeded into the lower chamber of a 24-well cell culture plate and allowed to adhere overnight. Activated CD8⁺ T cells were cultured in RPMI 1640 (Gibco) with 10% FBS (Gibco) and 1% penicillin–streptomycin, and 1 × 10⁵ CD8⁺ T cells per well were seeded into 0.4-μm pore-size transwell inserts with a 24-mm diameter (Corning, 3450) placed above the HFL1 monolayer. A 0.4-μm pore size was chosen to prevent cellular migration between compartments while allowing the diffusion of soluble factors. After 40 h of co-culture, CD8⁺ T cells were removed and fibroblasts were collected for subsequent experiments.

### PBMC and HFL1 co-culture assay

For PBMC–fibroblast co-culture experiments, HFL1 cells were seeded and maintained in the lower chamber as described above. Freshly isolated PBMCs were resuspended in RPMI 1640 (Gibco) supplemented with 10% FBS (Gibco) and 1% penicillin–streptomycin and pre-stimulated with recombinant human IL-2 (10 ng/mL). PBMC number and viability/proliferative capacity were evaluated by BrdU incorporation using flow cytometry, and only preparations with adequate viability were used for co-culture. 2 × 10^6^ PBMCs were then added to 0.4-μm pore-size transwell inserts (24-well format, 24-mm diameter; Corning, 3450) placed above the HFL1 monolayer. After 4 h of non-contact co-culture, bosutinib (10 μM) or TEW-7197 (10 μM) was added to the upper chambers, and the cultures were maintained for an additional 36 h. At the end of co-culture, HFL1 cells and/or PBMCs and supernatants were collected for subsequent analyses as indicated.

### Flow cytometry and Ki67 expression analysis

PBMCs were separated from peripheral blood of healthy controls and IPF patients for flow cytometry analysis by density gradient centrifugation using Ficoll-Hypaque (TBD science, LDS1075). PBMCs were washed by PBS containing 2% FBS and then fixed by 4% PFA. For intracellular cytokine staining and Ki67 staining, 1 × 10^6^ cells were permeabilized with Permeabilization Wash Buffer (Yeasen, 40403ES64) and stained with fluorescently-conjugated antibodies for 30 mins at room temperature according to manufacturer’s protocols. For surface staining, 1 × 10^6^ cells were stained with fluorescently-conjugated antibodies for 30 mins at room temperature. Staining antibodies for flow cytometry analysis include FITC-anti-CD3 (Thermofisher, 11-0037-41), PE-anti-CD8 (Thermofisher, 12-0088-42), anti-GZMK (Abcam, ab288726), IgG H&L (Alexa Fluor**®** 647, Abcam, ab150075) and Alexa Fluor™ 488-anti-Ki67 (Thermofisher, 53-5698-80). After washed with PBS, stained cells were recorded on Cytoflex (Beckman Coulter) and further analyzed by FlowJo_V10 software.

### Bleomycin-induced pulmonary fibrosis mouse model

To induce pulmonary fibrosis, 8–10-week-old male C57BL/6 mice (20–25 g) were anesthetized and administered bleomycin sulfate (5 mg/kg body weight) dissolved in sterile PBS via intranasal instillation. Control mice received an equal volume of sterile PBS. Mice were monitored daily for signs of distress or weight loss. Tissues and blood were harvested at day 21 post-BLM for histological and molecular analysis. In selected experiments, bosutinib (3.3 or 6.6 mg/kg) was administered to inhibit GZMK activity, and TEW-7197 (vactosertib), a dual TGFβR1/ALK4 inhibitor, was given orally every three days to block downstream signalling. Lungs and blood were collected at day 21 for histological, molecular and biochemical analyses.

### ELISA assay

The levels of GZMK in serum of bleomycin-induced IPF mice were analyzed using mouse GZMK (Hengrui Hongchuang Technology Development Co., China) ELISA kits according to the manufacturer’s instructions. The absorbance was read at 450 nm, and the results are presented as nanogram (ng) per liter (L).

### RNA extraction and real-time PCR

Total RNA was extracted from cultured cells or tissue samples using the RNAiso Plus reagent (Takara, 9108) according to the manufacturer’s instructions. The concentration and purity of RNA were determined using a NanoDrop spectrophotometer (Thermo Scientific). Complementary DNA (cDNA) was synthesized using a PrimeScript™ RT reagent Kit with gDNA Eraser (Takara, RR047A) in a total volume of 20 µL. Quantitative real-time PCR (qRT-PCR) was performed using TB Green® Premix Ex Taq™ II (Takara, RR820A) on a LightCycler® 480 System (Roche). Gene expression levels were normalized to GAPDH and calculated using the 2^^−ΔΔCt^ method.

### Western Blotting

Cells or tissue samples were lysed in RIPA buffer (Beyotime, P0013B) supplemented with protease and phosphatase inhibitor cocktails (Roche). Protein concentration was determined using the BCA Protein Assay Kit (Thermo Fisher). Equal amounts of protein (20–40 μg) were resolved by SDS-PAGE (10–12% gels) and transferred onto PVDF membranes (Millipore). Membranes were blocked with 5% non-fat milk in TBST (Tris-buffered saline with 0.1% Tween-20) for 1 hour at room temperature, followed by overnight incubation at 4°C with primary antibodies. After washing, membranes were incubated with HRP-conjugated secondary antibodies for 1 hour at room temperature. Signals were detected using ECL Western blotting substrate (Bio-Rad) and visualized using a ChemiDoc Imaging System (Bio-Rad). Densitometric analysis was performed using ImageJ software. Primary antibodies include anti-GZMK (CSB, PA002797), anti-a-SMA (proteintich, 67735-1-Ig), anti-collagen I (Abcam, ab260043).

### Hematoxylin and Eosin (H&E) Staining

Lung tissues were fixed in 4% paraformaldehyde for 48 hours, embedded in paraffin, and sectioned at 3 μm thickness. Sections were deparaffinized, rehydrated, and stained with hematoxylin for 5 minutes, followed by eosin staining for 2 minutes. After dehydration in graded ethanol and clearing in xylene, slides were mounted with neutral resin and examined under a light microscope (Olympus).

### Masson’s Trichrome Staining

Paraffin-embedded tissue sections were deparaffinized, rehydrated, and subjected to Bouin’s fixative at 56°C for 1 hour. Sections were stained sequentially with Weigert’s iron hematoxylin, Biebrich scarlet-acid fuchsin, phosphomolybdic-phosphotungstic acid, and aniline blue. Slides were dehydrated, cleared in xylene, and mounted. Collagen fibers stained blue, nuclei black, and cytoplasm red. Fibrosis severity was semi-quantitatively evaluated using the Ashcroft scoring system by two independent blinded observers.

### Immunohistochemistry (IHC)

Paraffin-embedded lung tissue sections were deparaffinized and rehydrated. Antigen retrieval was performed using sodium citrate buffer (pH 6.0) in a microwave for 24 minutes. Endogenous peroxidase activity was quenched with 3% hydrogen peroxide for 10 minutes. Sections were blocked with 5% normal goat serum for 30 minutes at room temperature, then incubated with primary antibodies overnight at 4°C. After PBS washes, sections were incubated with HRP-conjugated secondary antibodies for 30 minutes at room temperature. DAB substrate was used for color development, and nuclei were counterstained with hematoxylin. Slides were dehydrated, mounted, and imaged using a brightfield microscope. The following primary antibodies were used: anti-GZMK (CSB, PA002797), anti-ALK4 (abclonal, A2279).

### Multiplexed immunofluorescence staining

Surgical specimens were immediately fixed in neutral buffered formalin for 48 hours, followed by standard dehydration and paraffin embedding procedures. Five-micrometer sections were cut from paraffin blocks and mounted on positively charged glass slides. Prior to staining, sections underwent deparaffinization by baking at 70°C for 1 hour, followed by xylene treatment and rehydration through a graded ethanol series (100%, 85%, and 75%). Antigen retrieval was performed using EDTA buffer (pH 9.0) under optimized microwave conditions. To minimize non-specific binding, sections were blocked with 10% normal goat serum for 30 minutes at room temperature. Primary antibodies were applied and incubated overnight at 4°C in a humidified chamber. The following primary antibodies were used: anti-CD8 (Abcam, ab237709), anti-GZMK (Cell Signaling Technology, 98946), and anti-HSP70 (Proteintech, 10995-1-AP). Following PBS washes, sections were incubated with horseradish peroxidase-conjugated secondary antibodies according to manufacturers’ recommendations. Nuclear counterstaining was performed using DAPI.

### Statistical Analysis

All statistical analyses were performed using GraphPad Prism (version 9.5.0) and R (version 4.4.0). For comparisons between two groups, statistical significance was determined using either two-tailed Student’s t-tests (for normally distributed data) or Wilcoxon rank-sum tests (for non-parametric data). Correlation between continuous variables were evaluated using Pearson’s correlation coefficient for Spearman correlation analysis. For multiple group comparisons, one-way ANOVA was performed followed by Tukey’s post-hoc test to correct for multiple testing. Although normal distribution was assumed for certain analyses, formal tests of normality were not conducted due to sample size limitations. Statistical significance was defined as P < 0.05 for all analyses. Detailed statistical methods for specific comparisons are provided in the corresponding figure legends.

